# An interdisciplinary outpatient therapy program for children and adolescents with headache - real world data

**DOI:** 10.1101/2022.01.27.21268082

**Authors:** H Sobe, M Richter, R Berner, M von der Hagen, A Hähner, I Röder, T Koch, R Sabatowski, A Klimova, G Gossrau

**Affiliations:** Pain Center, University Hospital and Faculty of Medicine Carl Gustav Carus, TU Dresden, Germany; Department of Pediatrics, University Hospital and Faculty of Medicine Carl Gustav Carus, TU Dresden, Germany; Abteilung Neuropädiatrie, Medizinische Fakultät Carl Gustav Carus, Technische Universität Dresden; Smell & Taste Clinic, Department of Otorhinolaryngology, TU Dresden, Dresden, Germany; NCT Partner Site Dresden, Institute for Medical Informatics and Biometrics, Faculty of Medicine Carl Gustav Carus, TU Dresden, Germany; Department of Anesthesiology and Intensive Care, University Hospital and Faculty of Medicine Carl Gustav Carus, TU Dresden, Germany

**Keywords:** headache, children, therapy program

## Abstract

**Background/ Objective:** More than 2/3 of children and adolescents in Germany regularly suffer from headaches. Headache-related limitations in everyday life, school drop-out and educational impairment are common. Structured therapy programs for young headache patients are widely missing. We investigate the effects of an outpatient interdisciplinary headache program for children, adolescents and their parents.

**Methods:** 91 patients suffering from frequent headaches were treated in a 15 hour group program. Parents received 7 hours of therapy. At the beginning of the therapy program (T0), 6 (T1) and 12 months (T2) after completion, data on headache related disability (PedMidas), headache frequency, intensity, and pediatric pain disability score (PPDI) were collected. The primary endpoint was reduction in headache frequency, secondary endpoint reduction in PedMidas.

**Results:** 75 children and adolescents (9-18 years, median = 14; 66.7% female) and their parents provided prospective patient reported outcome measures. Patients were diagnosed with any form of migraine or tension type headache or a combination of both. 6 and 12 months after completion of the therapy program patients reported reduced headache frequency (headache days in the last three months median at baseline: 30; T1: 18 days; T2: 13 days). Linear mixed models revealed significant reduction over time (T0/T1 p = 0,002; T0/T2 p = 0,001). In addition, reduced headache disability has been reported 6 and 12 months after therapy (PedMidas median T0= 30, T1=15, T2=7; p<0,001 and p<0,001 respectively).

**Conclusions:** The interdisciplinary headache therapy program for children and adolescents, reported here, reduces headache frequency and headache related disability significantly in a period of 6-12 month following its completion. Comparative studies of children and adolescents with headaches in general outpatient treatment are needed to further describe the therapeutic gains of the program.

## Introduction

Evidence-based treatment options in young headache patients remain limited. Since headache is the most common reason for medical consultations and also the most common pain disorder in children and adolescents, there is the need to improve the situation(1). More than 2/3 of young people in Germany regularly suffer from headaches (2, 3). The majority of adolescent migraine patients continue to suffer from migraine in adulthood (4-6). Not least due to the lack of image morphological and functional correlates as well as biomarkers, headaches in children are often not perceived as a serious disease and diagnosis and therapy are not consistently pursued. Investigations in community samples depicted female gender, greater social impairment, depressive symptoms as negative prognostic factors in patients with frequent headaches in adolescents(7).

Young headache patients show high co-morbidity with further chronic pain (1, 8, 9). Lifestyle factors such as consumption of caffeine, lack of physical activity and psychosocial risk factors (e.g. family conflicts) are associated with frequent headaches in young patients (2, 5, 8, 10).

Approximately 5% of children and adolescents with headaches show severe headache-related limitations in everyday life from school drop-out to severe educational impairment (5, 11-13). Especially for children and adolescents with headache-related psychosocial restrictions, a therapy strategy that takes into account the biopsychosocial dimension of chronic headaches in their entirety is essential. Due to the changes that have already occurred in recurrent headaches, e.g. in behaviour, relaxation ability, physical activity, interdisciplinary therapy strategies are necessary, which include education, stress management, relaxation techniques and physical activation. Unimodal therapy strategies (e.g. exclusive drug treatment) in this complex situation are usually not enough (6, 14, 15).

Existing therapies for children and adolescents with headache include outpatient visits with pediatricians, who initiate additional diagnostics including cranial MRI, general laboratory exams and referral to ENT-specialists or ophthalmologists, acute analgesic prescription and occasionally referral for physiotherapy. Nevertheless, in the majority of patients this measures do not substantially change the frequency of headache. As discussed above, more intensive and individual therapy regimes are needed.

Therefore, we set out to develop an interdisciplinary outpatient therapy program, which includes headache education and a variety of practical therapies, enabling individual therapy approaches for young headache patients. Primary aim of the study is reporting therapy-induced change in headache frequency and headache-related disability 6 and 12 months after the therapeutic intervention.

## Methods

The study protocol was approved by the Ethics Board of the Faculty of Medicine at the TU Dresden (protocol number EK-462122017). Detailed information about the data collected were given to all participants and parents and informed written consent was obtained. All aspects of the study were performed in accordance with the Declaration of Helsinki.

### Patients

A cohort of 91 children and adolescents participated in the interdisciplinary outpatient therapy program with age-matched groups from January 2016 to June 2019. Patient data were prospectively collected at the start of the program, and after six and twelve months after its completion. The analysis in this manuscript is based on the data of 75 patients, who provided patient reported outcome measures for at least one follow up visit.

Therapy groups have been organized as a collaboration between Headache Outpatient Clinic, Pain Center and Department of Pediatrics, University of Dresden. All patients have been assessed by an interdisciplinary team of pediatrician, neurologist and pediatric psychotherapist.

Before participation in the program, patients underwent a detailed clinical examination and psychological evaluation. Motivated patients with a confirmed diagnosis of a primary headache disorder according to the ICHD[III criteria (16) for at least 6 months at the age of 9 to 18 years, headache-related limitations in school attendance, daily activities, quality of life were admitted to the therapy program.

### Interdisciplinary outpatient therapy program for children and adolescents

The interdisciplinary group therapy program consists of 8 therapy modules for patients. These include headache education, stress management, relaxation techniques, physical fitness, climbing therapy as communicative movement therapy to strengthen self-efficacy, art therapy, olfactory training as education and mindfulness training (Figure 1). Parent workshops with a focus on education take place on 4 parallel dates.

**Figure 1:**
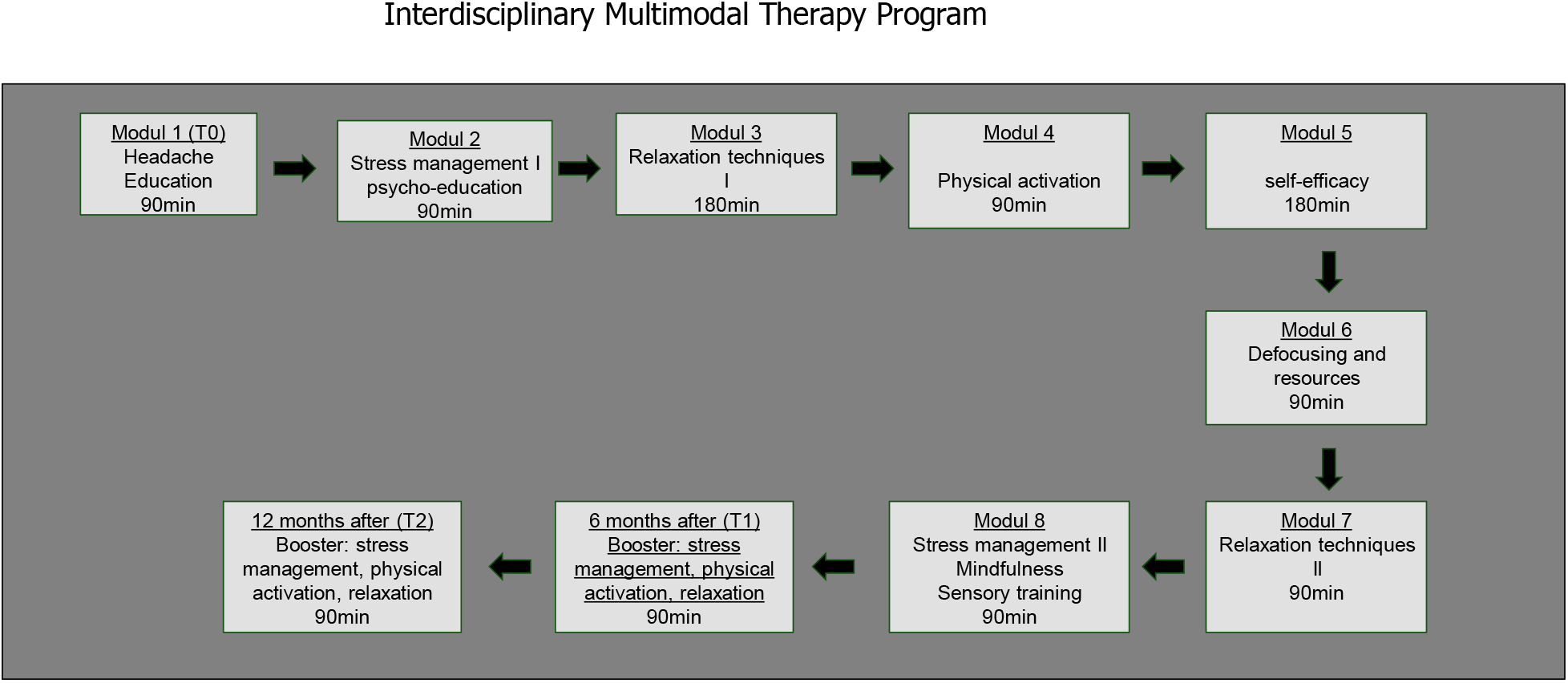
Therapy program overview

The therapy program is carried out over 2-3 months in afternoons or weekends and interuption of school is not necessary. The groups consist of 4-8 patients of one age group. Patients receive a total of 15 hours of therapy and their parents 7 hours. The aim of the program is to educate the patients about the disease and about non-drug and drug treatments in order to avoid missing or incorrect treatment. Recognition of stressors and possibilities of stress management are to be practiced, relaxation techniques and defocusing exercises are introduced and implemented in order to apply the individually appropriate technique in everyday life. Physical activation is encouraged as a team sport and on the climbing wall. Self-efficacy is developed centrally and the everyday transfer of new knowledge is stimulated. 6 and 12 months after the end of the program, group meetings are held. Here the current condition of the patients is reported, solution strategies for problems in implementing the therapy content are developed.

### Clinical data and Questionnaires

Clinical data as headache diagnosis, headache frequency and intensity, analgesic intake and concomitant diseases have been prospectively collected. In addition, two questionnaires, the pediatric migraine disability assessment score (pedMIDAS, range 0-240 points) (17) and pediatric pain disability score (PPDI) (18) have been used. The pedMIDAS is a validated questionnaire to quantify functional impairment due to headache over a period of 3 months. Sum scores >50 indicate severe, 31-50 moderate, 11-30 mild, 0-10 no or little impairment. The PPDI is a short questionnaire for self-or parent assessment of pain-related impairments in children and adolescents with chronic pain.

### Statistical procedures and data analysis

The data analysis was performed using SPSS vs. 29 (SPSS Inc., Chicago, Illinois). Categorical variables are summarized as frequencies and percentages; continuous variables are reported as means (M) ± standard deviations (SD) or medians (with interquartile range). For continuous variables, the independent samples or paired t-test, Mann-Whitney U test, Wilcoxon sign-rank test, F-test (ANOVA), or Kruskal-Wallis test were employed, as appropriate. For categorical variables, the association was assessed using the chi-squared test, the differences in distributions between time points were investigated using the McNemar–Bowker test. For hypothesis testing, p-values less than 0.05 (two-sided) were considered statistically significant. An adjustment for multiple comparison was performed when necessary. For mixed-effects models, p-values were obtained using the Satterthwaite’s method.

## Results

### Patients in the program

Descriptive statistics and further results are reported only for the sub-cohort of patients who consented to report outcome questionnaires for at least one follow up visit. This subcohort consists of 75 children and adolescents of ages 9-18 (median = 14, IQR=13:16), among whom 50 (66.7%) patients were female. The school type was available for 68 participants: 8 children (11.8%) attended primary school, 22 (32.4%) 6-year-secondary schools (secondary school level I certificates), 29 (42.6%) 8-year-secondary schools (university entrance level II certificates), 5 (7.4%) a vocational secondary school, 4 (5.9%) patients attended private or special schools. All of the participants had at least one primary headache diagnosis according to the IHS criteria (16). 16 patients (21.3%) were diagnosed with episodic migraine without aura (eMwoA), 9 (12%), with episodic migraine with aura (eMwA), 8 (10.7%) with chronic migraine (cM), 15 (20%) with episodic tension type headache (eTTH) and 3 (4%) with chronic tension type headache (cTTH), 9 (12%) with chronic tension type headache and episodic migraine (cTTH/eM) and 15 (20%) with episodic migraine and episodic tension type headache (eM/eTTH) (Figure 2).

**Figure 2:**
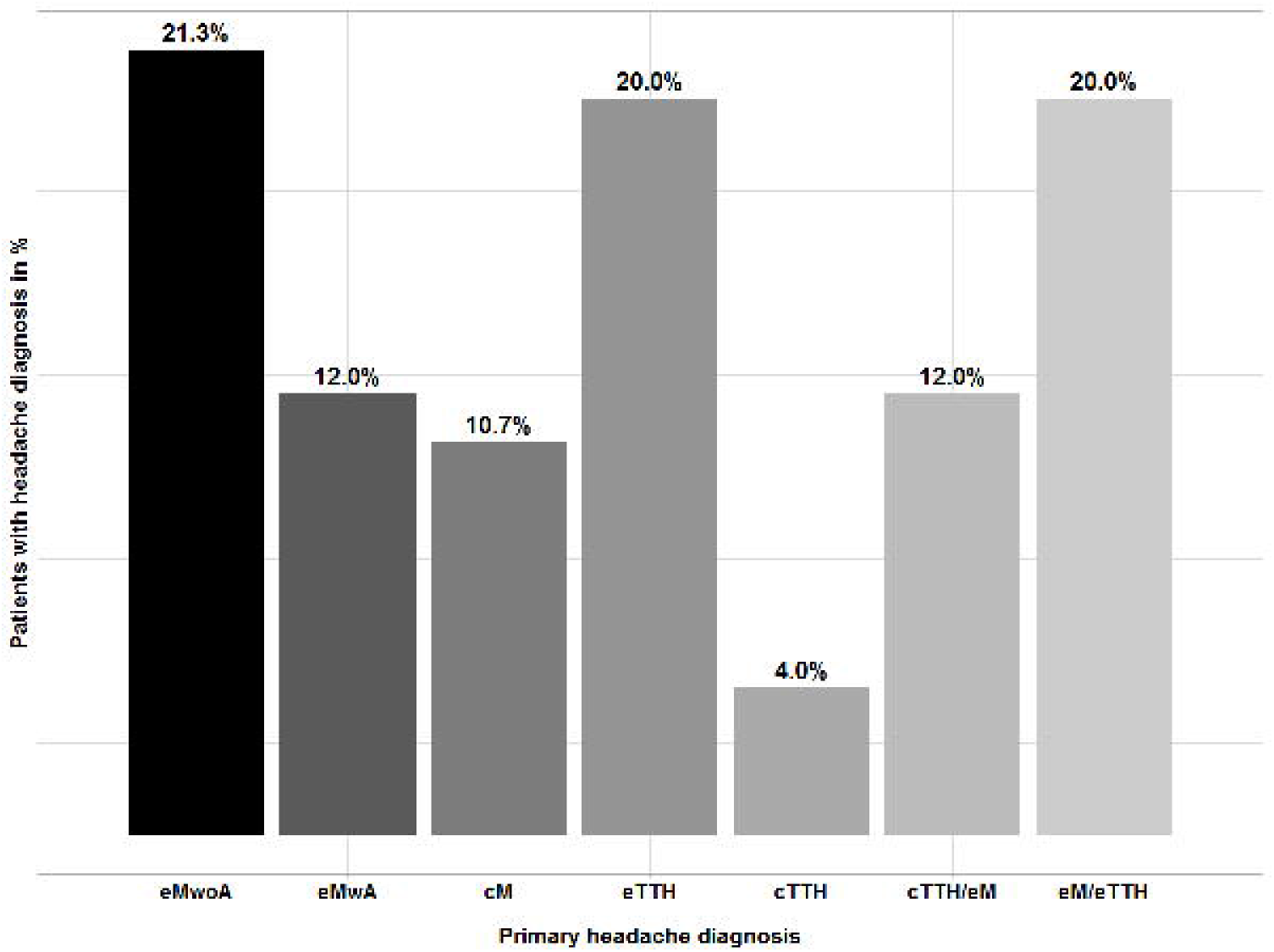
Primary headache diagnoses of patients in the program

In 45 (60.8%) participants, at least one accompanying disease was diagnosed. 18 (24.3%) patients suffered from back pain or abdominal pain, 9 (12.1%) had a neurological disease, 11 (14.9%) a mental disorder, 4 (5.4%) an endocrinological disorder and 18 (29.0%) with other diagnoses, i.e. dermatological or ophthalmological disease (Table 1).

**Table 1:** Frequency of additional diseases → this are new numbers already

|  | patients (n) | % of all patients |
| --- | --- | --- |
| Patients with concomitant diseases | 39 | 62,9 |
| other pain syndromes | 11 | 17,7 |
| chronic diseases | 32 | 51,6 |
| organic brain disorders | 6 | 9,7 |
| mental disorders | 14 | 22,6 |
| endocrinological diseases | 5 | 8,1 |
| other | 18 | 29 |

### Headache days at baseline and after group therapy

At the beginning of the program, headache frequency data were available for 72 participants (96%). Patients reported having had on average 43.74 (±33.44) headache days within the last three months, with the median of 30 days (IQR=15:90). Girls presented more headache days than boys: mean values 48.4 (± 34.3) and 34.9 (±30.5), medians 40 (IQR=15:90) and 20 (IQR= 15:40) days, respectively (W = 706, p = 0.158). Younger participants (up to the age of 13) showed on average 22 days less headache in the last 3 months than patients ≥ 14 years (W = 312, p = 0.008).

At the initial evaluation, the number of headache days differed significantly depending on the headache diagnosis (Kruskal-Wallis test, χ^2^ (3) = 16.194, p = 0.001). Patients with episodic migraine had significantly less headache days than those with chronic migraine (W = 36, p-value = 0.020).

Six months after the program, 65 (86.7%) patients provided information about their headache days during the last three months. The mean of 35.65(±34.19) days was reported (median = 18), with girls presenting on average 20.7 more headache days than boys (W = 635.5, p = 0.024). The overall difference between headache diagnoses was still considerable (Kruskal-Wallis test, χ^2^(3) = 17.096, p < 0.001), but the difference between patients with chronic and episodic migraine was not significant anymore (W = 51, p-value = 0.112).

Twelve months after completing the program, data were provided by 47 (62.7%) patients. The number of headache days in the last three month was on average 27.7 (± 30.9), with the median of 13, and thus was overall reduced compared to baseline and to 6 months after the program (Figure 3). No major differences between headache diagnoses were observed (Kruskal-Wallis, χ^2^ (3) = 6.595, p = 0.086).

**Figure 3:**
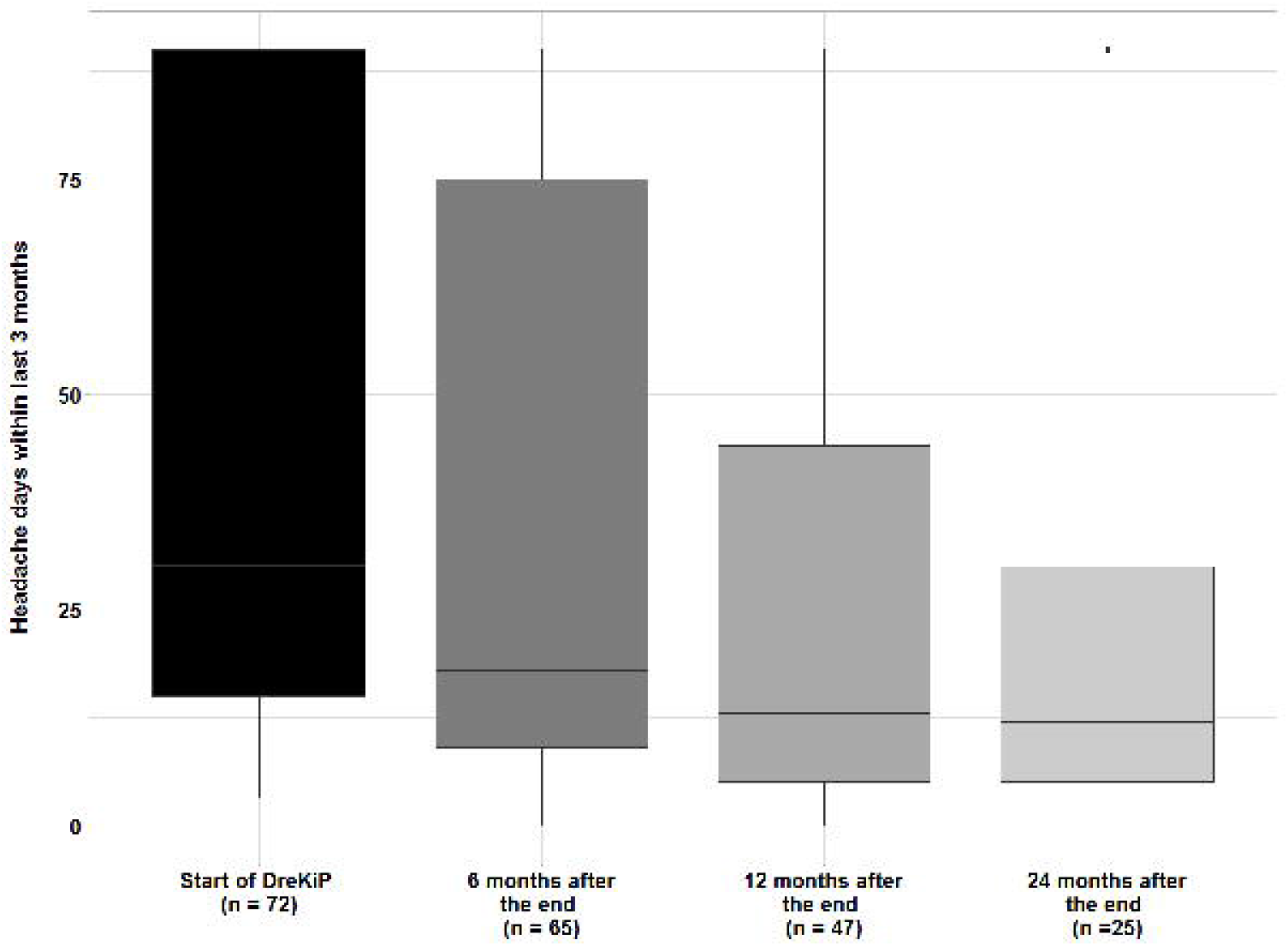
Headache frequency (Mean headache days ±SD in the last 3 months) at baseline and follow up 6, 12 and 24 months after the program

The two-year follow-up visit was completed by 25 (33.3%) patients. They had, on average, 23.32 (± 26.04) days, median = 12, which may indicate that the reduction achieved by 12 months remained stable. No significant differences between headache diagnoses were observed (Kruskal-Wallis, χ^2^ (3) = 0.145, p = 0.986).

Time trends in headache days were also investigated using a linear mixed-effects model with random intercept per patient and fixed effects for gender, time point, and main diagnosis. This analysis arrived to similar conclusions, indicating a strong reduction in headache days. As compared to the baseline, the average number of headache days decreased on average by 11.2 days per last 3 months after 6 month (p = 0.002) and by 17.3 days per last 3 months after 12 months (p < 0.001). Finally, after two-years, patients experienced on average 16.7 days less per last 3 months than at the baseline (p = 0.001). The interaction between time and gender was found borderline significant (χ^2^(3) = 7.975, p = 0.047). In particular, the estimated number of headache days of girls after 6 months was about 16.8 days per last 3 months more than those of boys at the same time point (p = 0.029). However, but no difference could be inferred after one year (p = 0.057) or two years (p =0.526).

### Headache intensity at baseline and after group therapy

At the beginning of the program, 69 (92%) patients provided data describing their headache intensity. Seven of them (10.1%) reported having mild, 30 (43.5%) moderate, and 32 (46.4%) severe headache, and the intensity seems to be associated with headache diagnosis (χ^2^(6) = 15.499, p = 0.017). While nobody reported having pain of a highest severity, patients with episodic migraine experienced severe headache more frequently than other patients (OR = 3.70, p = 0.026).

The intensity data of 57 (76%) patients were collected at the 6 months follow-up visit. Among them, one adolescent (1.7%) did no longer complain about headache, 13 patients (22.8%) had mild, 24 (42.1%) moderate, 15 (26.3%) severe headache, and four (7.0%) reported having the most severe headache. No association with headache diagnosis could be concluded (χ^2^(12) = 12.152, p = 0.434), and the difference in intensity between boys and girls was not significant (χ^2^(4) = 8.183, p = 0.085).

12 months after the program, the intensity information was obtained from 36 (48%) patients. One of them (2.8%) had no headache, 7 (19.4%) had mild, 17 (47.2%) moderate, 10 (27.8%) severe, and one patient (2.8%) the most severe headache (Figure 4). No difference due to headache type (χ^2^(12) = 17.135, p = 0.145) or to gender (χ^2^(4) = 7.722, p = 0.102) could be inferred.

**Figure 4:**
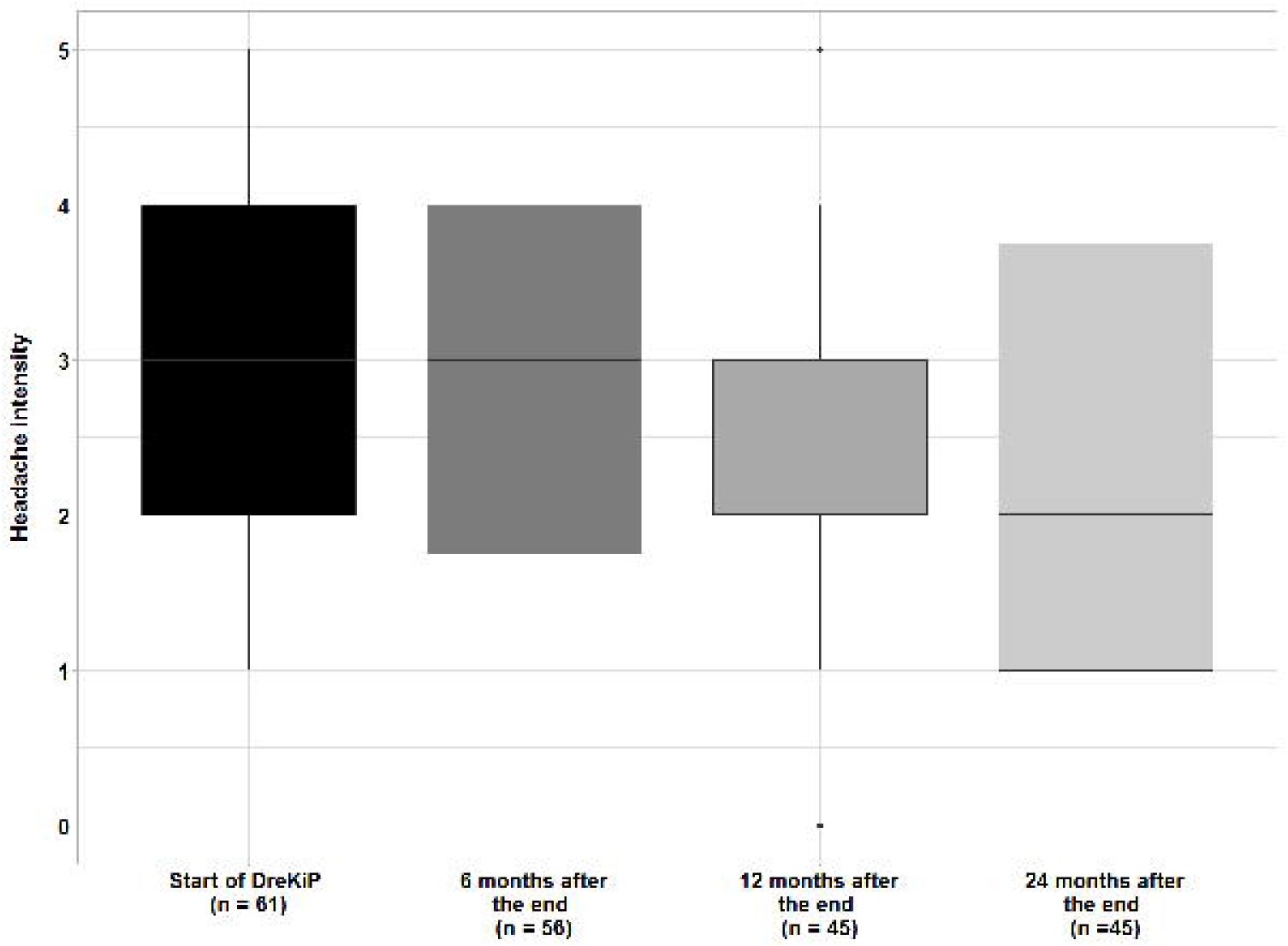
Average headache intensity (Mean headache intensity ±SD in the last 3 months) at baseline and follow up 6, 12 and 24 months after the program

Only 17 patients (22.7%) provided data on headache intensity at the two-year follow-up. Their data indicated no new trends as compared to the 12 months visit.

The effects of time, headache diagnosis, and gender on headache intensity were also explored using a linear mixed-effects model. As in the descriptive analysis, the effects of time and diagnosis were found to be significant (F = 4.089, p = 0.008, and F = 4.552, p = 0.006, respectively). A notable change in intensity was estimated to be after 12 months (about 30% decrease on average, p = 0.056). The estimated difference between baseline and 24 months was, on average, 40% (p = 0.025).

### Analgesic medication at baseline and after group therapy

Initially, the information about analgesic intake was provided by 72 patients (96%). Among them, 16 (22.2%) reported no analgesic medication for headache, 36 (50%) used analgesics more than once a month and 20 (27.8%) more than once a week.

Six months after the end of the program, the medication-intake data were available for 65 (86.7%) participants, of whom 19 (29.2%) children and adolescents did not need analgesics, 36 (56.9%) used medication more than once a month, and 9 (13.8%) more than once a week.

Twelve months after the end of the program, only 45 patients (60%) updated their information on analgesics. Out of them, 18 (40%) stated that they were not taking any medication, 20 (44.4%) reported taking it more than once a month, and 7 (15.6%) more than once a week (Figure 5). Finally, after two years, the data on analgesics were provided by 23 patients (30.7%). Only two of them (8.7%) were using analgesics more than once a week, while 11 patients (47.8%) reported that they were taking none.

**Figure 5:**
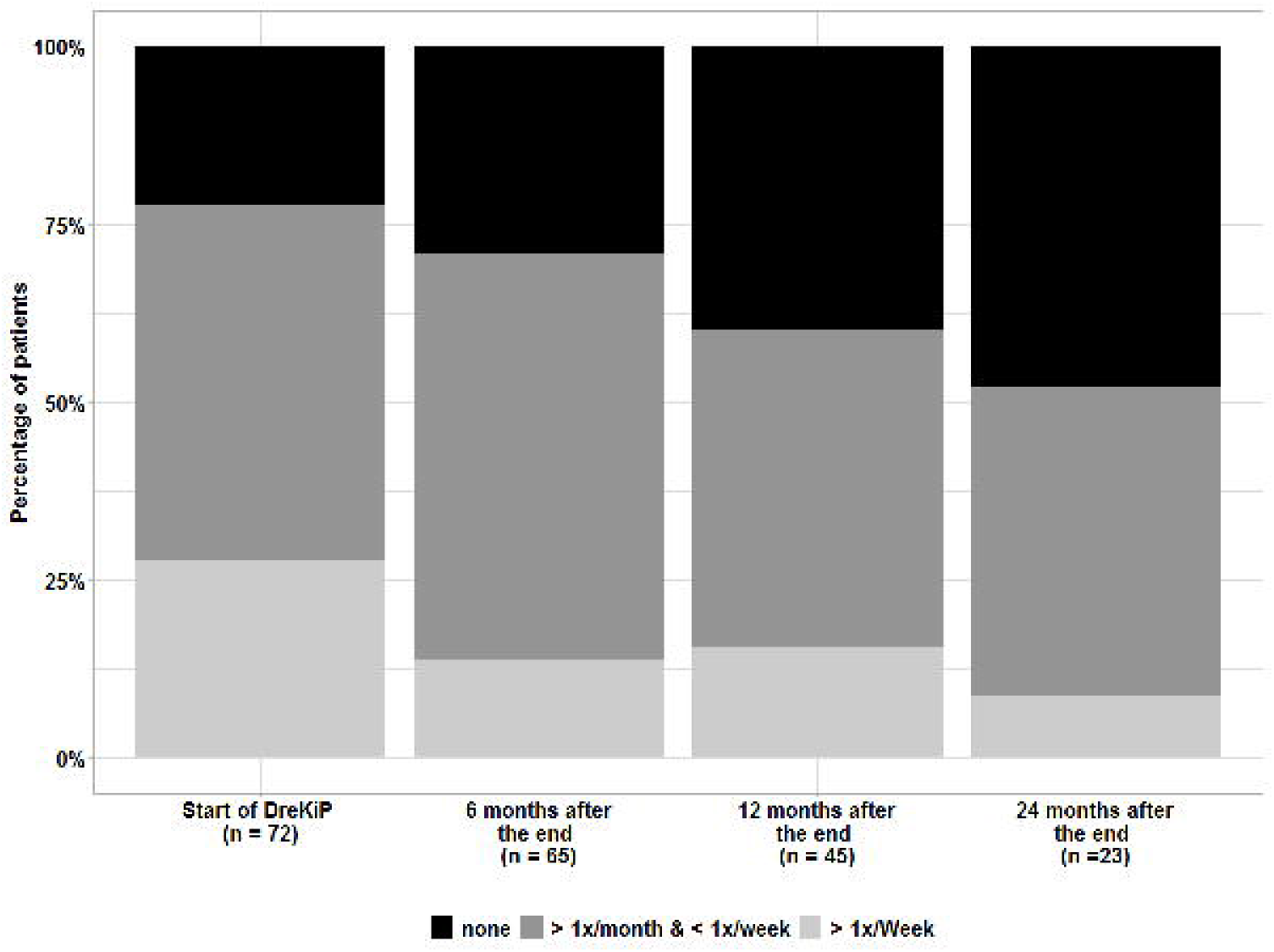
Acute Headache Medication intake at baseline and follow up, 6, 12 and 24 months after the program

No statistically significant changes in analgesic intake, compared to the baseline, were found at either 6 months or 12 months time points (McNemar–Bowker symmetry test, p = 0.105, p = 0.128, respectively), however, the two-year difference to baseline was found significant (p < 0.001). The association of analgesic intake with headache diagnosis and with gender at either time points was not significant.

### Headache related disability at baseline and after group therapy

At the beginning of the program, PedMIDAS scores as measure of headache related diability of 75 children and adolescents were obtained. The scores ranged from 1 to 200, with the mean of 40.51 (±39.25) and median of 30, indicating the presence of outliers. Patients with chronic migraine presented higher than average scores, with the mean of 66.0 (±13.81). The second largest mean score of 40.0 (±7.97) was observed for the episodic migraine group. However, no overall association with headache diagnosis could be inferred (F(3,70) = 1.69, p = 0.177, log-scale).

Six months after the program, PedMIDAS scores were obtained for 65 (86.7%) patients. The mean score 23.83 (± 24.93) was significantly lower than at the beginning of the program (V = 1465, p < 0.001). The maximum score of 120 and the median of 15 indicated a positive development as well. The difference in PedMIDAS due to headache diagnoses was not significant (F(3,61) = 1.167, p = 0.330, log-scale). As before, the chronic migraine group had the largest mean score of 36.6 (± 8.53), compared to other diagnosis groups.

The twelve-months data revealed a further reduction in PedMIDAS scores. The scores of 47 patients (62.7%) who returned for the follow-up ranged from 0 to 77 (median = 7), and their mean 16.89 (± 20.35), was significantly lower than the baseline value (V = 947.5, p < 0.001; Figure 6). As before, no overall association with headache diagnosis was found (F(3,43) = 1.987, p = 0.130, log-scale). The mean score of participants with chronic migraine was 41.6 (± 8.51).

**Figure 6:**
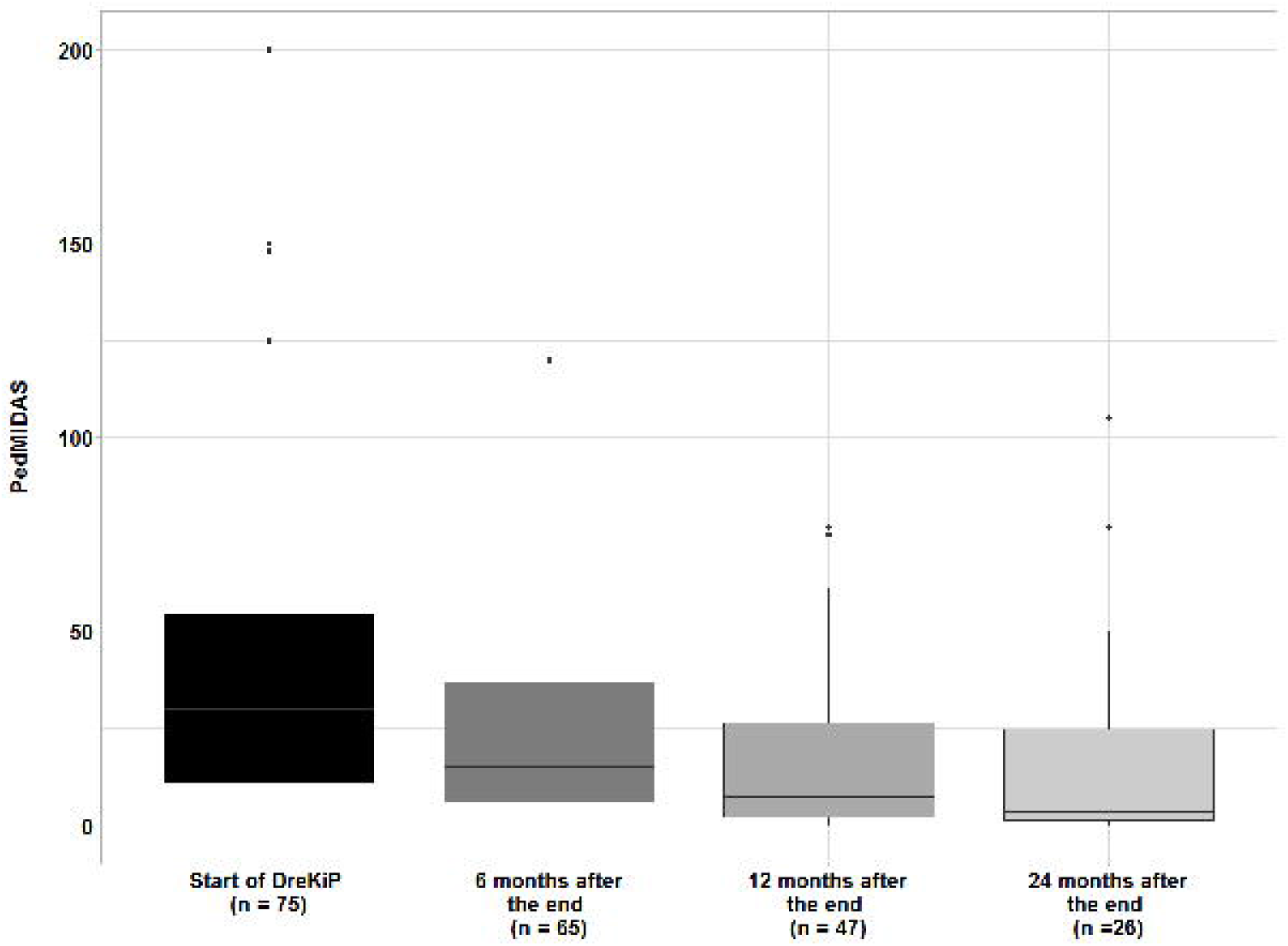
PedMidas (Mean±SD) at baseline and follow up 6, 12 and 24 months after the program

After two-years, PedMIDAS scores were obtained for 26 patients (34.7%). By then, the maximum score was 105, the mean was equal to 15.96 (± 26.16) and median = 3.5. Although these scores were not significantly different from the 12 months data (V = 89.5, p = 0.277), the improvement, compared to the start of the program, stayed significant (V = 296, p < 0.001). As in the previous time points, no association with headache diagnosis could be inferred (F(3,22) = 1.196, p = 0.334, log-scale), with the mean score of participants with chronic migraine being the highest, 30.8 (± 10.04).

Further, the percentage change in PedMIDAS score over time was investigated using a linear mixed-effects model. According to the fit results, the scores went down by 50% already in 6 months (p < 0.001), a decrease of about 70% was estimated to be achieved in 12 months (p < 0.001), and of almost 80 % in two years (p < 0.001). Across all time points, the estimated scores for girls were on average about 90% higher than for boys (p = 0.024).

### Pain related disability *at baseline and after group therapy*

At the beginning of the program, PPDI scores as measure of pain related disability were available for 63 (84%) children and adolescents. The minimum and maximum scores were 18 and 57, respectively, with mean = 33.13 (± 8.82), and median = 34.

Six months after the end of the program, PPDI scores were obtained from 60 (80%) patients. This time, the scores ranged from 12 to 57, with the mean of 29.93 (± 10.85) and median of 30.

The twelve-months scores were reported for 44 patients (58.7%). Although the score range stayed about the same, from 12 to 55, the new mean value of 27.91 (± 11.15) and median of 26 reflected a tendency of somewhat reduced pain disability (Figure 7).

**Figure 7:**
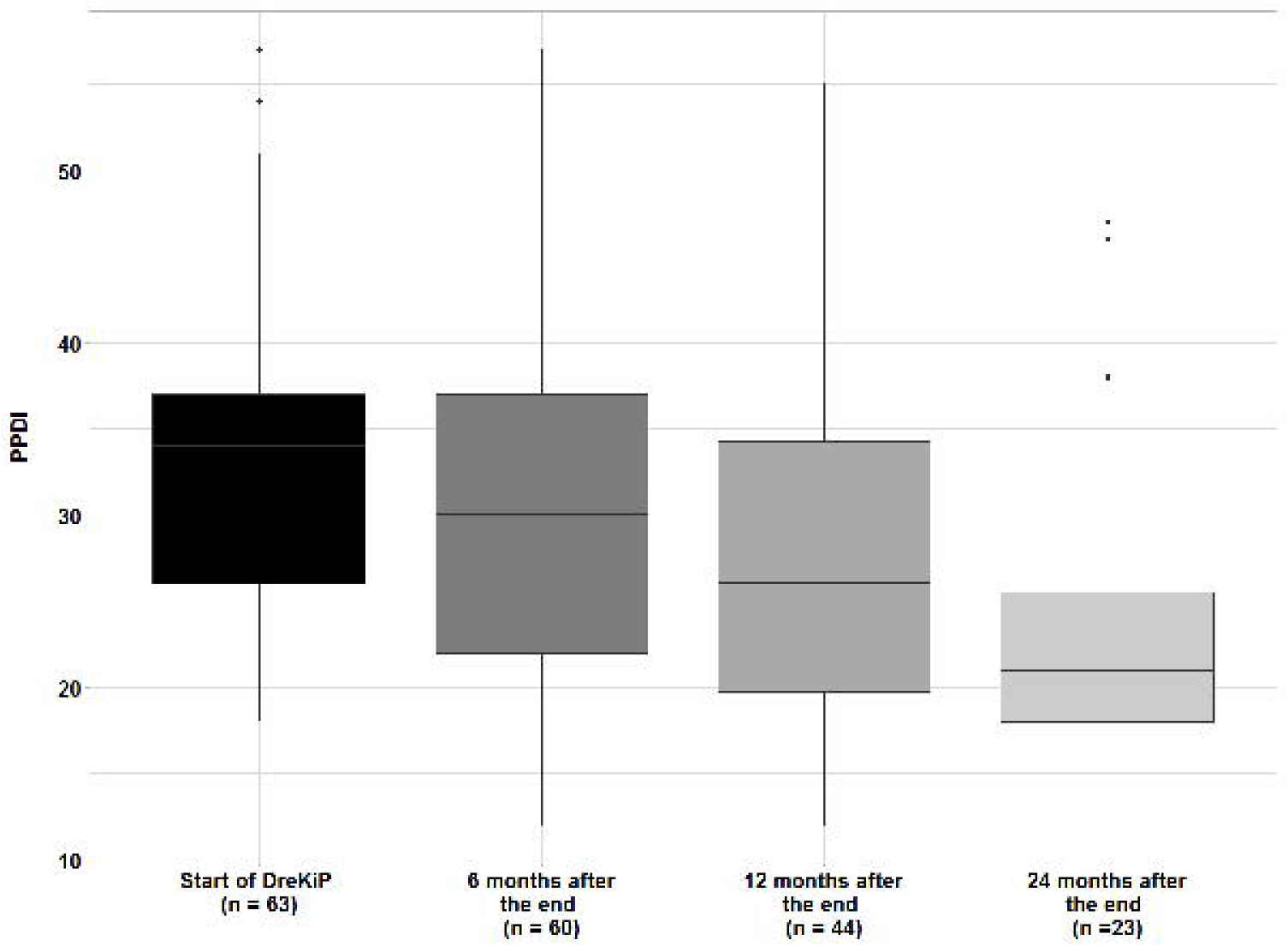
PPDI (Mean±SD) at baseline and follow up 6, 12 and 24 months after the program

The two-year data were collected from 23 patients (30.7%). The scores, ranged from 12 to 47, with the mean of 23.61 (± 9.63) and median of 21 indicated a small reduction in pain disability (Figure 7). At either time point, no significant differences in PPDI scores due to the headache diagnosis were found.

An additional analysis with linear mixed-effects models indicated an overall significant improvement in PPDI by about 25% starting from baseline to 24 months (p = 0.001), but implied a slightly different time dynamic in PPDI scores of boys and girls (χ^2^(3) = 7.864, p = 0.049). Throughout the whole time, girls’ scores were estimated to be about 20% higher than those of boys (p = 0.022). By six months, boys’ scores decreased by 25% (p = 0.001), while girls’ scores stayed, on average, about the same (p = 0.3961). According to the model, a gradual decrease continued for both genders, leading to almost a 30% reduction in girls (p < 0.001) and 20% reduction in boys (p = 0.076) in 24 months after the program.

## Discussion

Our results indicate that the interdisciplinary multimodal group therapy program presented here is effective in improving headache-related limitation of daily functioning for adolescents with headache in a period of 6 to 12 months after the program.

61.3% of the patients were female, reflecting the more frequent occurrence of headaches in girls beginning with adolescence (7). Most patients in the program were older than 14 years. This reflects the epidemiology of primary headache in childhood and adolescence, with prevalence increasing with age (6). The majority of patients suffered from any type of migraine. Considering the psychosocial impairment that migraine has already in young patients, it seems compelling that these patients in particular have a need for intensified therapy (19).

62.9% of patients suffered from at least one other disease. Comorbidity of headaches and a range of diseases has been described in pediatric migraine (20). It includes psychiatric disorders, head and neck injuries, cardiovascular disease, metabolic syndrome, asthma, sleep apnea and other pain syndromes and points towards multimorbidity beginning early in life. In adults all classes of comorbidities significantly increased the risk of progression from episodic to chronic migraine (21). Children and adolescents with concomitant diseases also suffer more frequently from headaches (2). Thus, it is obvious that a large proportion of patients treated in the program here, show an increased risk for headache chronification.

In adult patients with migraine, interdisciplinary multimodal treatment programs have been shown to effectively reduce monthly headache days and improve daily function (22-24). Unimodal or bimodal interdisciplinary treatment programs have been associated with improvement in headache frequency in children and adolescents (25-28). However, there is limited data on the treatment effects of interdisciplinary treatment programs for children and adolescents with headaches (19). Especially, there are only a few prospective studies published, mainly investigating all types of pain in children and adolescents (29-31). One study evaluated the effectiveness of interdisciplinary pain treatment in children with chronic headache (30). 50 patients aged 10-19 years, 62% female attended an interdisciplinary therapy program 8 h/day, 5 times/week for 2-7 weeks. After the program, disability measured by Headache Impact Test-6 reduced significantly from severe impact at baseline to some impact at 6-8 week follow-up. Disability and school participation improved. Another inpatient interdisciplinary treatment program for pain has been investigated using a waiting control group, each 52 patients (29). Significantly more children in the treatment program compared to those in the waiting group improved in the combined end point improvement, consisting of pain intensity, disability, school absence directly after the program.

Compared to the sparse data available from other therapy programs, our patient population shows significant improvements in daily function 6 and 12 months after the end of the program. This is accompanied by a significant reduction in the number of headache days 6 and 12 months after the end of the program. This is also reflected in the reduced pain disability after the program. However, it should be noted that the PPDI is valid as a measure of chronic pain impairment in children (32) but does not capture the extent of impairment as well as the PedMidas, especially for episodic headache patients. However, since 17,7% of the children also reported concomitant pain other than headache, the PPDI can also describe the extent of the overall development. For the chronification of migraine, a connection with comorbid pain has been described in adults (33). The programme described here, can have additional effects due to the versatile therapy content. Further studies are required to make a precise statement on this.

### Strengths and limitations

To our knowledge, this is the first prospective evaluation of a multimodal interdisciplinary outpatient program for children and adolescents with recurrent or chronic primary headaches. Headache related disability, headache frequency and overall pain disability were significantly reduced 6 and 12 months after the program. The 15 hour group therapy program for children and adolescents with 7 hours parents therapy showed to be effective in improving headache related disability and headache frequency.

However, our study has several limitations. First of all, there is a number of patients with incomplete or missing data. Furthermore, we studied a mixed group of headache patients, although chronic tension-type headache and episodic migraine might have different pathophysiologies. Due to the small number of cases per diagnosis group, further evaluations are not possible. More patients should be studied after completion of the programme in the future. Furthermore, we see gender-specific differences in the headache related disability. This point has not been taken into account in the current therapy contents and should be considered and further investigated in the future.

### Conclusions

Our study demonstrates that interdisciplinary group therapies are a valuable treatment for children and adolescents with frequent and limiting headaches. Further establishment and investigation of the effectiveness of these therapies should be encouraged. Not least because relevant social and economic disease burdens arise already in childhood and adolescence due to frequent headaches.

### Bullet Points

#### Clinical implications

- Outpatient interdisciplinary headache treatment program for children and adolescents with headache effectively reduces headache-related limitations
- effects persist up to one year, indicating an prophylactic long term effect

#### Public health implications

- Awareness of headache in children and adolescents should be improved and preventive measures addressed
- Interdisciplinary therapy programs should be implemented in routine care of children and adolescents with headaches

## Data Availability

All data produced in the present study are available upon reasonable request to the authors.

## Acknowledgements

We thank the participants of this investigation.

The interdisciplinary treatment program received funds from the “Dresdner Kinderhilfe” (https://www.dresdner-kinderhilfe.de), a non-profit association for the support of children with chronic diseases.

The authors disclose no conflict of interest.

## Author contributions

G.G., M.R. Study concept and design. H.S., A.K.: Acquisition of data. H.S., A.K., I.R., G.G.: Analysis and interpretation of data. M.R., G.G., A.H. R.B., T.K., M.v. H., R.S.: Drafting and critical revision of the manuscript.

## Legends for Illustrations and tables

Table 1: Frequency of additional diseases (multiple answers possible)

## Notes

### Competing Interest Statement

The authors have declared no competing interest.

### Clinical Trial

DRKS00027523 Deutsches Register klinischer Studien, Germany

